# Readiness to start long acting injectable HIV treatment and prevention and conditions for acceptance among clinic clients in Uganda

**DOI:** 10.64898/2026.10.14.26365499

**Authors:** Stephen Charles Olet, Wandera Uthmaan Muluga, Drake Musoki, Bernard Raymond Kihumuro, Farid Muluga, Ivan Ssebandeke, Fiona Among, Chad D. Stecher, David Musoke

## Abstract

Long-acting injectable human immunodeficiency virus (HIV) medicines expand treatment and prevention choices, but willingness to use them does not establish readiness to begin. We examined readiness to start these options, associated factors and conditions for acceptance among people living with HIV and pre-exposure prophylaxis (PrEP)-service clients in Uganda.

This cross-sectional study analysed 225 participants at Mildmay Hospital and Kajjansi Health Centre IV in central Uganda. Readiness meant preferring initiation immediately or within three months. Modified Poisson regression estimated adjusted prevalence ratios (aPRs) and 95% confidence intervals (CIs), including age-by-service-group interaction. Separate models examined informational, perceptual and financial factors, adjusting for service group, age and sex. Logistic marginal standardisation checked demographic and expanded age comparisons.

Overall, 108 participants (48.0%) were ready to start injectable HIV treatment or prevention, 111 (49.3%) wanted more information about these options, and six (2.7%) preferred initiation in six to twelve months. Readiness was 80.8% among people living with HIV aged 15–24 years and 34.4% among those aged 25 years or older (aPR 2.35, 95% CI 1.63–3.38). Corresponding proportions among PrEP-service clients were 39.2% and 41.4% (aPR 0.95, 95% CI 0.60–1.50; interaction p=0.002). In separate adjusted models, clinic-staff information (aPR 1.94, 95% CI 1.42–2.66), belief in greater effectiveness (2.00, 1.43–2.80) and a one-point increase in benefit score (3.27, 2.36–4.55) were associated with higher readiness. A one-point increase in concern score (0.70, 0.60–0.80) and financial difficulty sometimes or more often (0.68, 0.48–0.96) were associated with lower readiness.

Fewer side effects was the leading condition for accepting injectable HIV treatment or prevention across service and readiness groups. Nearly half of participants were ready to start, while a similar proportion wanted further information about injectable options. Programmes should assess intended initiation timing alongside willingness and address safety, appointment flexibility, privacy and affordability through counselling and service planning.

## Introduction

Long-acting injectable medicines expand choices for human immunodeficiency virus (HIV) treatment and prevention, but their public-health benefit depends on informed decisions and sustained access. Monthly cabotegravir and rilpivirine maintained viral suppression without inferior virological outcomes compared with oral antiretroviral therapy in a phase 3 maintenance-treatment trial [1]. For prevention, injectable cabotegravir reduced HIV acquisition compared with daily oral tenofovir disoproxil fumarate and emtricitabine in the HIV Prevention Trials Network (HPTN) 083 and 084 trials [2,3]. Treatment and pre-exposure prophylaxis (PrEP) serve different clinical purposes and have different eligibility requirements. Both introduce decisions about replacing daily tablets with scheduled injections and their associated service demands.

In Uganda, injectable options would enter services where clients weigh treatment or prevention benefits against privacy, affordability and the practical demands of continued care. Qualitative studies among Ugandan people living with HIV and women receiving services to prevent vertical HIV transmission describe reduced pill burden and greater privacy alongside concerns about adverse effects, supply and affordability [4,5]. Experience in rural Uganda and Kenya also demonstrates the importance of delivery arrangements: a randomised trial extension reported mean biomedical prevention coverage of 69.7% with a flexible package including injectable cabotegravir, compared with 13.3% under standard care [6]. This result reflects the combined package of product choice and service delivery, rather than the injectable medicine alone.

Ugandan surveys have documented willingness to use injectable PrEP among 65.8% of 234 clients at a Kampala prevention clinic and 69.8% of 285 adolescent girls and young women in Kampala [7,8]. These findings establish interest, but do not show when clients would prefer to begin or which conditions they would want met before accepting injectable HIV treatment or prevention. Willingness describes interest in an option, whereas preferred initiation timing distinguishes near-term intention from a decision deferred for information or other reasons. Neither measure establishes clinical eligibility or actual uptake.

We examined readiness to start long-acting injectable HIV treatment or prevention among clinic clients in Uganda. We described preferred initiation timing, compared readiness by age within treatment and prevention groups, examined informational, perceptual and financial correlates, and summarised conditions for acceptance and reasons for reluctance to use injectable options.

## Materials and methods

### Study design and setting

We conducted this primary cross-sectional study within the quantitative component of a mixed-methods research project at Mildmay Hospital and Kajjansi Health Centre IV in Wakiso District, central Uganda, from 4 May to 31 July 2026. Mildmay is a private, not-for-profit hospital providing specialised HIV care, treatment, training and research alongside other clinical services. Kajjansi Health Centre IV is a public-sector facility providing routine HIV prevention and treatment within primary healthcare. The two facilities offered complementary service contexts: a specialised HIV centre and a public facility serving surrounding communities. Both provide services for people living with HIV and clients accessing PrEP. This manuscript reports the client survey, comprising people living with HIV and PrEP-service clients, rather than the healthcare-provider or qualitative components of the project. Mildmay’s public facility profile reports direct care and treatment for more than 15,000 people living with HIV and non-HIV services for more than 11,000 people. These published service counts describe the facility’s scope and are not study-period recruitment denominators[9].

### Participants and analytical cohort

Participants were aged at least 15 years and belonged to the HIV-treatment or PrEP-service group. We used age-stratified recruitment from facility registers, followed by random selection within strata. People who were too ill to participate or had communication difficulties precluding participation were excluded. The prevention group included current, former and never users of oral PrEP. We therefore use the term PrEP-service clients rather than describing all prevention respondents as current PrEP users.

The parent sample-size calculation targeted a difference in willingness between the two service groups, assuming proportions of 74% and 89%, a two-sided 5% significance level, 80% power and equal allocation. The protocol estimated approximately 104 respondents per group and increased the total for anticipated non-response to 232 participants, or 116 per group. This calculation was for the parent willingness question, not for readiness, interaction tests or the present exploratory models.

The raw export contained 251 records, comprising 131 people living with HIV and 120 PrEP-service clients. We restricted this analysis to the exact participant identifiers retained in the parent primary analysis: 116 in each service group. Thus, 15 HIV-treatment records and four prevention records were outside the selected cohort. The same four PrEP participants excluded from the parent analysis were excluded here, and no new sample was drawn. Cohort restriction ensured comparability with related manuscripts but should not be interpreted as evidence that surplus records were clinically ineligible. The prevention cohort comprised 55 current, 33 former and 28 never users of oral PrEP. The saved roster, rather than a fresh random selection, defined inclusion.

Thirty-two selected PrEP-service clients answered “No” to receiving care at either named facility. We retained these respondents in the primary analysis to reproduce the parent cohort and assessed restriction to respondents answering “Yes” as a sensitivity analysis.

### Survey measures

We used a study-specific structured questionnaire developed to address the research objectives. It covered sociodemographic circumstances, treatment or prevention experience, prior awareness, information sources, effectiveness beliefs, willingness, preferred initiation timing, conditions for acceptance, and perceived benefits and concerns. Trained research assistants administered the questionnaire electronically using Open Data Kit, with English and Luganda participant materials. The questionnaire elicited views about injectable HIV options. It did not record prescriptions, dispensing, injection administration, subsequent clinic attendance or product-specific clinical eligibility.

The primary outcome was stated readiness, derived from the preferred timeline to switch or start. “As soon as possible” and “within three months” were coded ready; “six to twelve months” and “after more information” were coded not yet ready. “Not applicable” was excluded from the primary binary outcome but retained in the full timing distribution. Waiting for information expresses an unresolved decision rather than a calendar-based delay. We therefore interpret the binary classification as intended near-term initiation, not a validated multidimensional readiness scale. The strict sensitivity definition counted only “as soon as possible” as ready.

Service group distinguished people living with HIV from PrEP-service clients. Age was categorised as 15–24 or at least 25 years, consistent with the parent protocol, and sex was included in demographic adjustment. Willingness was classified as willing or very willing versus neutral, unwilling or very unwilling. We used this measure to describe its relationship with readiness and to restrict one sensitivity analysis, rather than making willingness the principal outcome.

Clinic-staff information was indicated by reporting clinic staff as an information source, while other information sources could also be selected. The effectiveness-belief indicator distinguished “more effective” from less effective, equally effective or uncertain responses. Awareness distinguished understanding the basics or being well informed from having heard little or not having heard of injectable options. Financial distress concerned financial difficulty meeting basic needs during the previous three months. We compared “sometimes”, “often” or “always” with “never” or “rarely”. Although the output label is “frequent financial distress”, this category includes “sometimes” and is described accordingly in the paper.

Perceived benefit was the mean of six five-point items: reduced pill burden, improved privacy or reduced stigma, better adherence, convenience during travel, feeling normal, and overall benefit versus oral medication. Responses ranged from strongly disagree to strongly agree. Perceived concern, labelled risk score in the analysis output, was the mean of eight four-point items: pain or discomfort at the injection site, injection-site reactions, side effects lasting longer than oral ART, strict clinic attendance requirements, cold-chain or drug availability concerns, eligibility requirements, long-term safety unknowns, and overall risk versus oral medication. Response labels were not concerned, neutral, concerned and highly concerned. Higher scores indicated greater benefit or concern, respectively. A complete set of item responses was required for each mean, and the selected cohort had no missing score values. These are questionnaire-derived summaries of perceptions, not validated clinical risk measures. Effect estimates refer to a one-point increase on their respective mean-score scales.

Conditions for accepting injectable HIV options and reasons for reluctance allowed multiple responses. Acceptance-condition options were fewer side effects, flexible appointments, transport support, privacy, provider recommendation and other. Reasons for reluctance were analysed among neutral or unwilling respondents and covered injection fear, side effects, long-term safety, clinic frequency, privacy or stigma, cost or transport, preference for oral medication and other. Percentages used respondents in the relevant subgroup as denominators and need not total 100%. Free-text review identified 25 explicit “None” entries among 26 other acceptance-condition responses and one request for free treatment. We interpreted explicit “None” as no additional condition specified, without treating missing text as equivalent. Subgroup findings for the other code are not relabelled as no-condition responses because the available subgroup table does not separately classify its free text.

### Statistical analysis

We described cohort flow, initiation timing, readiness and multiple-response items using counts and percentages. The primary readiness analysis included 225 participants with applicable timing responses. We fitted modified Poisson regression with a log link and heteroskedasticity-consistent (HC0) sandwich standard errors to estimate adjusted prevalence ratios (aPRs) and 95% confidence intervals [10]. The demographic model included service group, age category and sex. An interaction model added age-by-service group and provided younger-versus-older contrasts within each service group. With this interaction present, the service-group coefficient applies to the older reference age group and is not an overall comparison between treatment and prevention clients.

Separate exploratory models added one proposed factor at a time to service group, age and sex. These models examined benefit score, concern score, clinic-staff information, effectiveness belief, awareness and financial distress. They did not mutually adjust all proposed factors or include the age-by-service interaction. The forest plot combines factor estimates from these separately adjusted models with the two age contrasts from the interaction model. It is therefore a display of related analyses, not the coefficient set of one fully adjusted model.

An expanded exploratory model included service group, age, their interaction, sex, benefit and concern scores, clinic-staff information and effectiveness belief. Awareness and financial distress were not included in this expanded model. Some modified Poisson fitted means exceeded one, especially in the expanded model. We therefore used logistic regression and marginal standardization to report expanded-model age contrasts and checked a corresponding demographic interaction model using the same approach. For each service-age combination, standardization averaged predicted probabilities over the observed covariate distribution of the 225-person analytical cohort. Younger-versus-older prevalence ratios were calculated from these probabilities. Percentile confidence intervals used 500 bootstrap resamples. Expanded estimates condition on contemporaneously measured perceptions and information, so they are not interpreted as total causal age effects or formal mediation results.

Sensitivity analyses restricted the cohort to participants willing to use injectable HIV treatment or prevention, used only immediate initiation as ready, counted not-applicable responses as not ready, and restricted participants to those reporting care at either named facility. We also cross-tabulated readiness by age, interview date, form version and device among people living with HIV. These recruitment checks examined whether observed patterns occurred within shared versions or devices; they did not identify interviewers or resolve unmeasured session differences. All analyses were unweighted, used available complete cases, and were performed with the accompanying R script. Exploratory confidence intervals and p-values were not adjusted for multiple comparisons and should be interpreted as supporting investigation rather than confirming independent determinants.

### Ethics

Mildmay Uganda Research Ethics Committee approved the study (MUREC-2026-1828). Adults provided written informed consent before participation. For participants aged 15–17 years, a parent or guardian provided written consent and the adolescent provided assent. Participation was voluntary, and interviews took place in private. Registration was also obtained from the Uganda National Council for Science and Technology (UNCST).

## Results

### Cohort and preferred timing

The analysis retained 232 participants from the 251-record export, with 116 in each service group. The HIV-treatment group contained 52 participants aged 15–24 and 64 aged at least 25 years, while the prevention group contained 55 and 61 participants respectively. All seven not-applicable timing responses occurred among prevention clients, leaving 51 younger and 58 older prevention clients in the primary readiness analysis. Age, sex and the proposed model factors had no missing values in the selected cohort. Table 1 presents the full timing distribution by age and service group.

**Table 1.** Preferred timing to start injectable HIV treatment or prevention by service group and age.

| Timing | PLHIV 15–24<br>n=52 | PLHIV 25+<br>n=64 | PrEP 15–24<br>n=55 | PrEP 25+<br>n=61 |
| --- | --- | --- | --- | --- |
| As soon as possible | 42 (80.8) | 18 (28.1) | 19 (34.5) | 23 (37.7) |
| Within three months | 0 (0.0) | 4 (6.2) | 1 (1.8) | 1 (1.6) |
| Six to twelve months | 0 (0.0) | 2 (3.1) | 2 (3.6) | 2 (3.3) |
| After more information | 10 (19.2) | 40 (62.5) | 29 (52.7) | 32 (52.5) |
| Not applicable | 0 (0.0) | 0 (0.0) | 4 (7.3) | 3 (4.9) |

Values are n (%), using the full selected service-age subgroup. PLHIV, people living with HIV; PrEP, pre-exposure prophylaxis. Not applicable is retained here but excluded from binary readiness models.

Among 225 participants with an applicable timing response, 108 (48.0%) were ready to start injectable HIV treatment or prevention. Of these, 102 preferred initiation immediately and six within three months. Of the 117 not yet ready to start, 111 wanted further information about injectable options and six preferred initiation in six to twelve months.

Readiness to start injectable treatment was reported by 42/52 (80.8%) younger and 22/64 (34.4%) older participants living with HIV. Among PrEP-service clients, readiness to start injectable prevention was 20/51 (39.2%) among younger and 24/58 (41.4%) among older respondents with applicable timing responses. All 42 ready younger participants living with HIV preferred immediate initiation. Fig 1 shows preferred initiation timing across the four service-age groups.

**Fig 1.**
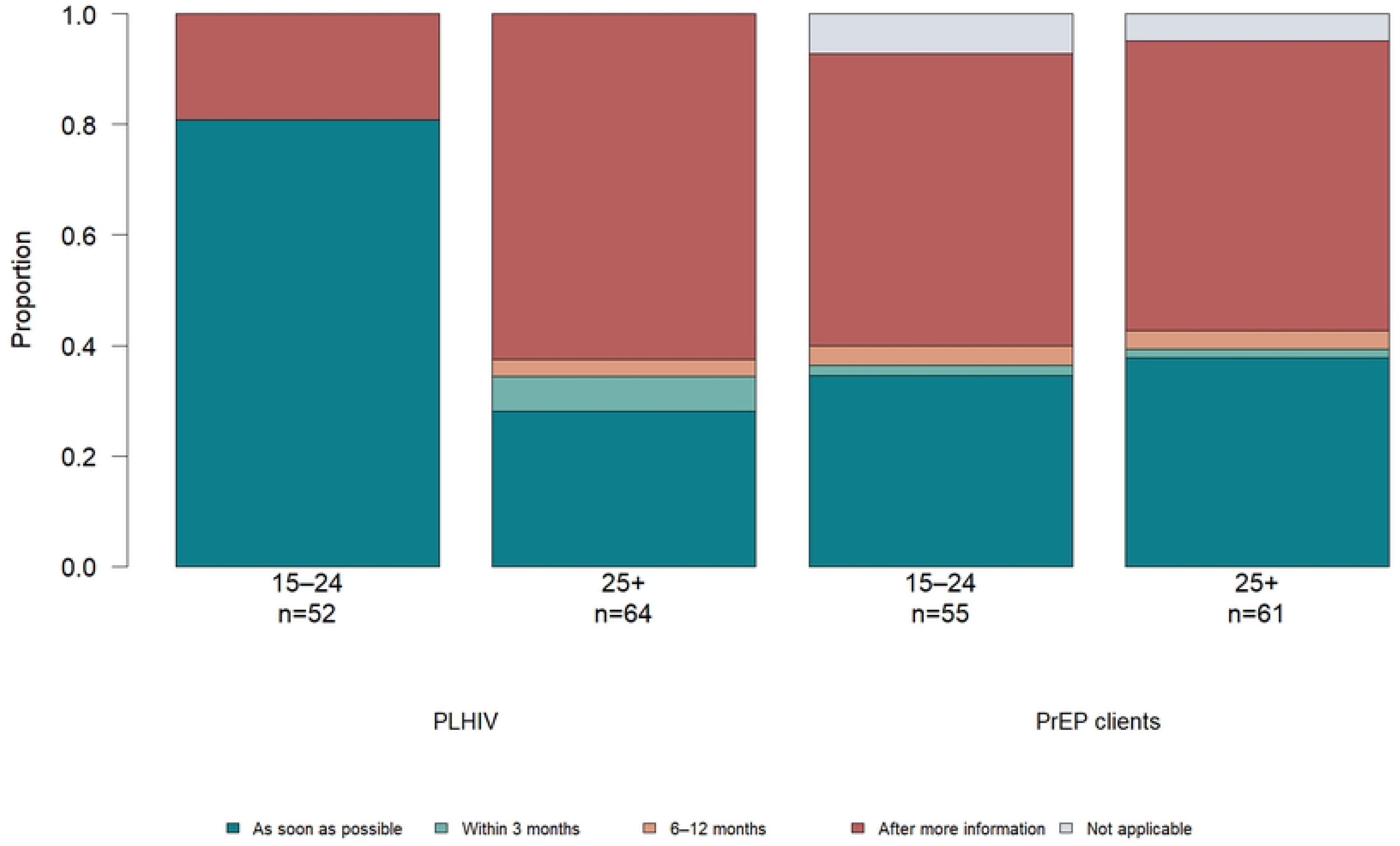
Preferred timing to start injectable HIV treatment or prevention by service group and age. Bars show the five timing categories for all 232 participants, including seven not-applicable responses among PrEP-service clients. Percentages use the full service-age subgroup as the denominator.

Willingness to use injectable HIV treatment or prevention and readiness to start did not coincide. Among 176 participants willing to use injectable HIV treatment or prevention with an applicable timing response, 99 (56.3%) were ready to start within three months and 77 (43.8%) were not yet ready. Nine of 49 neutral or unwilling participants with applicable responses were nevertheless ready to start within three months.

### Age and service group

In the demographic interaction model, younger participants living with HIV had higher readiness to start injectable treatment than older participants (aPR 2.35, 95% CI 1.63–3.38). There was no clear age difference in readiness to start injectable prevention among PrEP-service clients (aPR 0.95, 95% CI 0.60–1.50). The age-by-service interaction ratio was 0.40 (95% CI 0.22–0.72; p=0.002). Within the older age group, the PrEP-versus-HIV-treatment readiness contrast was 1.21 (95% CI 0.76–1.92).

Demographic logistic marginal standardisation gave closely similar age contrasts: 2.35 (95% CI 1.65–3.57) among people living with HIV and 0.95 (95% CI 0.60–1.44) among prevention clients. In the expanded standardised model, the age contrast among people living with HIV was smaller at 1.77 (95% CI 1.30–2.54), while that among prevention clients was 1.06 (95% CI 0.74–1.48). Table 2 distinguishes the two standardisation models and their predicted readiness prevalences.

**Table 2.** Logistic marginal standardisation by service group and age.

| Group or contrast | Demographic model | Expanded model |
| --- | --- | --- |
| PLHIV 25+ years | 34.34 (22.81–47.20) | 37.64 (27.07–48.07) |
| PLHIV 15–24 years | 80.71 (68.07–90.60) | 66.50 (52.53–83.41) |
| PrEP client 25+ years | 41.47 (28.94–54.44) | 45.13 (34.83–57.12) |
| PrEP client 15–24 years | 39.26 (25.28–51.72) | 48.02 (33.86–61.44) |
| PLHIV young vs older PR | 2.35 (1.65–3.57) | 1.77 (1.30–2.54) |
| PrEP client young vs older PR | 0.95 (0.60–1.44) | 1.06 (0.74–1.48) |

Values are standardised readiness percentages or younger-versus-older prevalence ratios, with 95% confidence intervals. Demographic adjustment includes service group, age, their interaction and sex. Expanded adjustment additionally includes benefit score, concern score, clinic-staff information and effectiveness belief. Both models use 225 participants.

### Information perceptions and financial circumstances

In separate demographic-adjusted models, reporting clinic staff as an information source was associated with higher readiness to start injectable HIV treatment or prevention (aPR 1.94, 95% CI 1.42–2.66). Believing injectable options were more effective than oral options was also associated with higher readiness (aPR 2.00, 95% CI 1.43–2.80). Each one-point increase in mean perceived benefit was associated with an aPR of 3.27 (95% CI 2.36–4.55), whereas each one-point increase in mean concern was associated with an aPR of 0.70 (95% CI 0.60–0.80). Financial difficulty sometimes or more often was associated with lower readiness than never or rarely experiencing difficulty (aPR 0.68, 95% CI 0.48–0.96). Understanding the basics or being well informed, compared with little or no awareness, showed no clear association (aPR 1.28, 95% CI 0.92–1.76). Fig 2 displays selected estimates; the complete regression outputs are presented in S1 Appendix.

**Fig 2.**
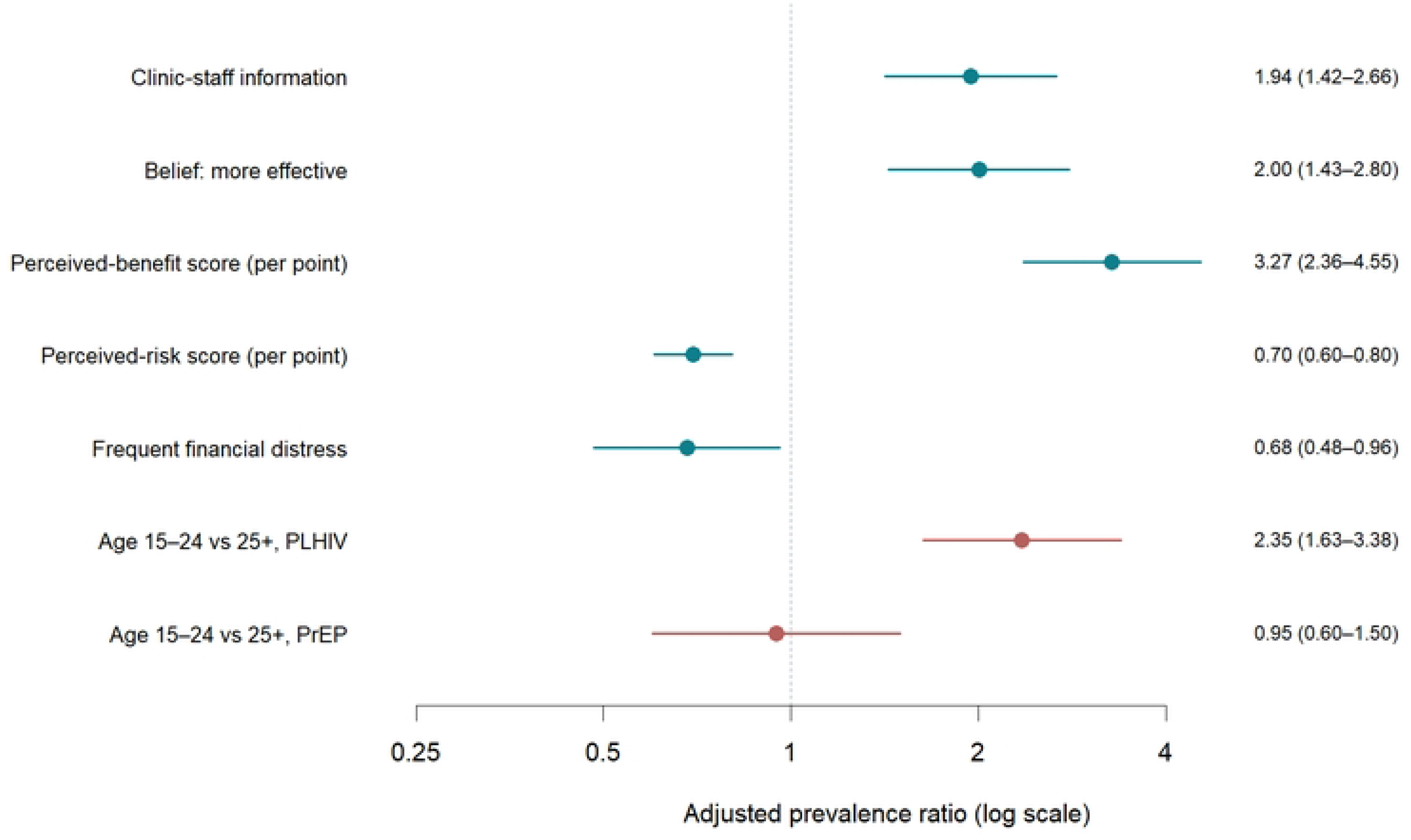
Adjusted prevalence ratios for readiness to start injectable HIV treatment or prevention from demographic and exploratory models. Clinic-staff information, effectiveness belief, benefit score, concern score and financial difficulty estimates each come from a separate model adjusted for service group, age and sex. Age contrasts come from the demographic model including age-by-service interaction and sex. These estimates are not mutually adjusted coefficients from one expanded model. Score estimates are per one-point increase; financial difficulty includes sometimes, often or always versus never or rarely. Horizontal lines indicate 95% confidence intervals.

The expanded modified Poisson model produced a maximum fitted mean of 1.61, whereas the corresponding logistic model had a maximum predicted probability of 0.99 (S1 Appendix). The expanded standardised age comparisons are presented in Table 2.

### Conditions for accepting injectable HIV options and reasons for reluctance

Fewer side effects was the most frequently mentioned named condition for accepting injectable HIV treatment or prevention in every service-readiness subgroup: 28/64 (43.8%) ready and 31/52 (59.6%) not-yet-ready participants living with HIV, and 27/44 (61.4%) ready and 36/65 (55.4%) not-yet-ready PrEP-service clients. Flexible appointments were mentioned by 9/64 (14.1%) ready and 11/52 (21.2%) not-yet-ready participants living with HIV, compared with 17/44 (38.6%) and 24/65 (36.9%) PrEP-service clients. Approximately three in ten respondents in each subgroup mentioned privacy. Fig 3 presents the five named conditions, with subgroup counts and percentages in S1 Appendix.

**Fig 3.**
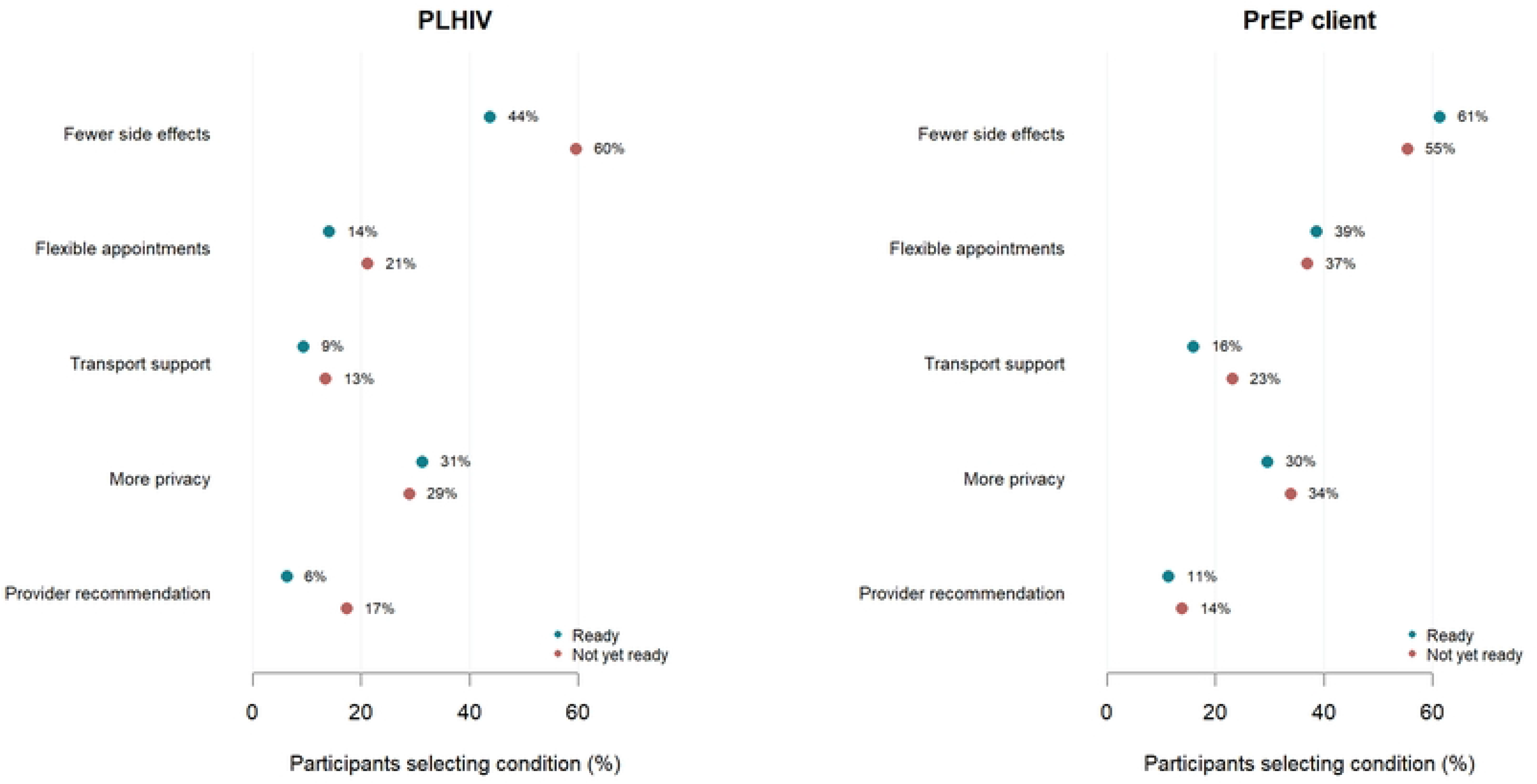
Conditions for accepting injectable HIV treatment or prevention by readiness and service group. Dots show the proportion mentioning each named multiple-response option. Denominators are 64 ready and 52 not-yet-ready people living with HIV, and 44 ready and 65 not-yet-ready PrEP-service clients. Percentages need not sum to 100%. Other free-text responses are described separately.

Transport support was mentioned by 6/64 (9.4%) ready and 7/52 (13.5%) not-yet-ready participants living with HIV and by 7/44 (15.9%) and 15/65 (23.1%) prevention clients. Provider recommendation was mentioned by 4/64 (6.3%) ready and 9/52 (17.3%) not-yet-ready participants living with HIV and 5/44 (11.4%) and 9/65 (13.8%) prevention clients. Among 26 participants selecting other across the full cohort, 25 explicitly wrote “None” and one requested free treatment.

Reasons for reluctance to use injectable HIV treatment or prevention were available for 55 neutral or unwilling respondents, comprising 12 people living with HIV and 43 PrEP-service clients. Side effects were mentioned by 8/12 (66.7%) and 24/43 (55.8%), respectively, fear of injections by 4/12 (33.3%) and 12/43 (27.9%), and long-term safety concerns by 1/12 (8.3%) and 13/43 (30.2%). Eight of 43 PrEP-service clients (18.6%) mentioned cost or transport concerns, compared with none of the 12 people living with HIV. Table 3 reports these subgroups descriptively, including respondents with not-applicable readiness responses.

**Table 3.** Reasons for reluctance to use injectable HIV treatment or prevention among neutral or unwilling participants.

| Reason | PLHIV n=12 | PrEP clients n=43 |
| --- | --- | --- |
| Fear of injections | 4 (33.3) | 12 (27.9) |
| Side effects | 8 (66.7) | 24 (55.8) |
| Concerns about long-term safety | 1 (8.3) | 13 (30.2) |
| Clinic visit frequency | 1 (8.3) | 7 (16.3) |
| Privacy or stigma concerns | 1 (8.3) | 5 (11.6) |
| Cost or transport challenges | 0 (0.0) | 8 (18.6) |
| Preference for oral medication | 0 (0.0) | 2 (4.7) |
| Other (specify) | 0 (0.0) | 3 (7.0) |

Values are n (%). Multiple selections were allowed. Includes six respondents with not-applicable timing; no stratification by readiness was performed.

### Sensitivity analyses and recruitment checks

Restricting the analysis to participants willing to use injectable HIV treatment or prevention with applicable timing responses retained a higher readiness prevalence among younger people living with HIV: aPR 2.30 (95% CI 1.58–3.34) in the interaction model. The interaction remained evident (ratio 0.37, 95% CI 0.21–0.66; p<0.001). The stricter immediate-initiation outcome identified 102/225 (45.3%) as ready, and counting not-applicable responses as not ready identified 108/232 (46.6%). Restriction to those reporting care at either named facility left 196 participants with applicable timing responses, of whom 99 (50.5%) were ready. The pooled younger-versus-older aPRs were 1.77 (95% CI 1.31–2.38) for immediate initiation, 1.59 (95% CI 1.20–2.10) when not applicable was coded not ready, and 1.82 (95% CI 1.38–2.40) in the care-restricted analysis (S1 Appendix).

Interview metadata showed no overlap in recruitment days between younger and older participants living with HIV. Younger respondents were interviewed from 31 May through 6 June, while older respondents were interviewed on dates from 24–30 May, with four additional interviews on 8 and 16 June. Version 3 of the electronic form was used for 27 younger and 60 older respondents. Within that version, readiness was 20/27 (74.1%) and 19/60 (31.7%), respectively. On a device used for both age groups, readiness was 28/37 (75.7%) among younger and 3/17 (17.6%) among older respondents. A second shared device recorded 14/15 (93.3%) and 8/24 (33.3%).

## Discussion

Almost half of respondents with applicable timing responses were ready to begin injectable HIV treatment or prevention, while a similar proportion wanted further information about these options. Readiness was higher among younger people living with HIV, whereas age differences were small among PrEP-service clients. Interest alone therefore provides an incomplete estimate of near-term demand. These findings complement Ugandan willingness studies and East African implementation evidence emphasising informed choice and flexible delivery [7,8,6]. For planning, clinics should assess intended timing and unresolved questions during counselling rather than use willingness alone to estimate immediate demand for injections. Aggregated preferences would help services plan assessment appointments, counselling capacity and follow-up while retaining options for clients who defer initiation.

Only 56.3% of participants willing to use injectable HIV treatment or prevention with applicable timing responses were ready to start injectable HIV treatment or prevention within three months. The willingness and readiness items therefore captured different aspects of the decision, although their occasional discordance also raises questions about how participants understood them. The 48.0% readiness estimate should not be interpreted as lower acceptability than the 65.8% willingness reported at a Kampala prevention clinic or the 69.8% reported among adolescent girls and young women in Kampala: those studies measured willingness, recruited different populations and did not use this timing classification [7,8]. Qualitative research also describes preferences evolving during discussion rather than existing as fixed commitments [11,12]. Asking about both interest and timing would help clinics distinguish those seeking an immediate assessment from those who need further explanation or time to decide.

Reporting clinic staff as an information source was associated with higher readiness in a separate demographic-adjusted model. Interested clients might have sought information from staff, and greater clinical engagement could explain both exposure and readiness; the data do not establish a counselling effect. Even so, the association is consistent with evidence that trusted providers help clients judge whether new treatment options suit them [13,14]. In Kampala, inadequate counselling and unclear referral communication contributed to delayed oral treatment initiation [15], while prevention clients in rural Eastern Africa valued respectful care and provider support [16]. These findings support attention to the content and accessibility of counselling. A practical service response is to ask what clients already know, explain treatment and prevention separately, and offer follow-up opportunities for questions rather than equating contact with staff with informed understanding.

Higher perceived benefit was associated with readiness to start injectable HIV treatment or prevention, while greater perceived concern was associated with lower readiness in separately adjusted models. Participants appear to weigh anticipated relief from daily tablets against what an injection and its service requirements could entail. Ugandan and Kenyan qualitative studies describe reduced pill burden, privacy and easier adherence alongside reservations about side effects and capacity to deliver injections [4,5,17]. Mobile men in Malawi also preferred injections for discretion and reduced risk of forgetting tablets, but raised concern about reactions interfering with work [18]. The substantial benefit-score ratio should nevertheless be interpreted on its actual scale: a one-point increase in a mean five-point score is large, and both the score and readiness were measured in the same interview. The association supports discussing personally relevant advantages and concerns, but does not quantify the improvement in readiness achievable through counselling.

Believing injectable options were more effective was associated with higher readiness to start injectable HIV treatment or prevention, whereas broad self-rated awareness showed no clear association. This suggests that familiarity and the meaning attached to a product are not interchangeable. Ugandan research among young people considering oral treatment similarly linked readiness with treatment beliefs, including beliefs whose clinical accuracy required attention [19]. Among West African men discussing injectable PrEP, perceived better protection coexisted with doubts about efficacy and safety [20]. Our effectiveness item also spans two different applications: injectable cabotegravir was superior to the oral comparators for HIV prevention in HPTN 083 and HPTN 084, whereas maintenance treatment with cabotegravir and rilpivirine was noninferior to continued oral treatment in the studied population [2,3,1]. Counselling should make those distinctions clear rather than suggesting that any injection is necessarily a more effective treatment for every client.

Fewer side effects remained the leading named condition for accepting injectable HIV treatment or prevention even among participants ready to begin. Readiness therefore coexisted with reservations rather than signalling unconditional acceptance. Ugandan studies describe fear of adverse effects and injection discomfort alongside enthusiasm for injectable treatment [4,5]. Women in the HPTN 084 qualitative substudy and clients who had used cabotegravir in Uganda and Kenya also weighed privacy and convenience against pain and access concerns [21,12]. These accounts distinguish anticipated fears from experiences after initiation; our survey addresses the former. Providers should explain the expected reactions and their management, allow clients to weigh alternatives, and establish how concerns would be reviewed after a first injection. The 25 explicit “None” free-text responses indicate no additional condition specified, but should not be treated as evidence that those respondents had no concerns elsewhere in the questionnaire.

Prevention clients selected flexible appointments more often than treatment clients in both readiness strata, while privacy was mentioned by roughly three in ten respondents across groups. These are descriptive comparisons rather than tested differences, but they point to service delivery as part of the product choice. In East African cabotegravir-user interviews, unpredictable work and travel sometimes complicated injections despite overall enthusiasm [12]. West African participants raised similar scheduling concerns [20], and formative pharmacy-delivery research in Kenya identified longer opening hours, rapid services and privacy as potential advantages [22]. The East African trial extension showed that a flexible package including injectable choice could substantially increase prevention coverage [6]. Together, these findings favour assessing locally feasible appointment and follow-up arrangements while preserving oral prevention options for clients whose circumstances or preferences make injections unsuitable.

Financial difficulty sometimes or more often was associated with lower readiness in a separate demographic-adjusted model. The question concerned meeting basic needs and does not identify the cost of injections, observed missed appointments or willingness to pay. Nevertheless, financial strain could make transport, time away from work or other service demands harder to absorb. Cost and accessibility were raised in the HPTN 084 qualitative substudy, while Ugandan women considering injectable treatment expressed concerns about affordability and supply [21,5]. Kenyan pharmacy-delivery stakeholders also identified clients’ ability to pay as a potential barrier [22]. These studies support assessing the full cost of accessing care, not merely the price of a medicine. Our financial estimate was not mutually adjusted for all perceptual factors, so prospective implementation studies should examine whether affordable access and practical support improve initiation among interested clients experiencing economic strain.

Younger people living with HIV were more often ready to start injectable treatment than older participants, whereas prevention clients showed no clear age difference. The service-specific pattern argues against treating age as a universal indicator of demand. Younger treatment clients could find relief from daily medication particularly attractive, but age-linked preferences vary across settings. For example, Kenyan adults and youth described both enthusiasm and safety reservations, while South African youth emphasised effectiveness and discretion in prevention choices [17,23]. Caregiver relationships also shape adolescents’ engagement with injectable-treatment research in Uganda and South Africa [24], and young people elsewhere report diverse views on needles, follow-up and safety information [25]. The public health implication of these findings is to assess young clients’ circumstances and preferences directly rather than assuming they will all favour injections or that older clients will not.

The observed age difference among people living with HIV persisted in the willing-only model and appeared within shared form versions and devices. However, younger and older treatment clients were interviewed on separate days, so their different recruitment contexts could explain part or all of the contrast. Shared-version and shared-device comparisons cannot resolve that problem. Ugandan evidence on oral treatment readiness and the adolescent injectable-treatment findings show why psychosocial and relational context belongs alongside chronological age when studying treatment decisions [19,24]. Until recruitment-session arrangements are clarified and the pattern is replicated with overlapping recruitment, the result should be presented as an association in these differently recruited age groups. It provides a question for subsequent investigation rather than a basis for prioritising one age group for access.

The younger-versus-older contrast among people living with HIV was smaller in the expanded logistic standardisation than in the demographic model. This does not prove that benefit perceptions or clinic information mediated the difference. The expanded estimate conditions on variables measured alongside readiness, some of which could precede the decision, result from it or share unmeasured causes. Studies of Ugandan injectable-treatment acceptability and provider communication describe closely connected beliefs, trust and preferences, making several explanations plausible [4,14]. The two estimates therefore answer different adjustment questions. The demographic standardised result closely agreed with the robust Poisson contrast, and the expanded result describes the comparison after accounting for selected perceptions and information. Neither establishes a causal pathway from age through beliefs to readiness.

Among respondents neutral or unwilling to use injectable HIV treatment or prevention, side effects and fear of injections were commonly mentioned. PrEP-service clients also mentioned long-term safety and cost or transport. These responses identify topics for counselling without establishing a single explanation for reluctance. Ugandan qualitative research among people living with HIV describes concern about injection safety and service capacity, while the HPTN 084 substudy and East African cabotegravir-user interviews show how privacy and convenience coexist with concern about pain and access [4,21,12]. Providers should distinguish fear of the injection procedure, expected short-term reactions and uncertainty about long-term safety because these concerns require different explanations and support. The 12-person treatment subgroup is too small for broader comparisons. Counselling should support an informed decision, preserve a suitable oral option when preferred, and avoid treating reluctance as disengagement from care.

The information-dependent responses, safety conditions and appointment preferences suggest programmes should plan a decision process alongside injection delivery. Clinics should ask about willingness to use injectable treatment or prevention and intended timing, identify unresolved questions and practical requirements, then undertake product-specific assessment for clients wishing to proceed. East African experience with flexible prevention choice supports offering suitable oral and injectable options through responsive services, while Ugandan treatment research highlights counselling and clear navigation of care [6,16,15,4]. Services should explain appointment schedules, expected reactions, follow-up and available support before clients decide, and offer further counselling when requested. A request for information should prompt support rather than classification as rejection. Prospective evaluation should document information received, actual initiation, reasons for deferral, appointment attendance and continued use. Linking these outcomes with affordability and delivery arrangements would show whether services help interested clients act on their preferences.

### Strengths and limitations

A strength of this primary study is its distinction between willingness to use injectable HIV medicines and intended initiation timing, which identifies unresolved decisions and service requirements among potential users. It addresses a different question from the related willingness and oral PrEP-history manuscripts. The analysis used the same participant roster, retained the full timing distribution, examined alternative readiness definitions and audited recruitment metadata. Multiple-response conditions added service detail, and logistic marginal standardisation checked age comparisons using bounded probabilities. These strengths do not establish clinical eligibility or observed uptake. Readiness to switch treatment should not delay clinically indicated oral therapy, and readiness for an injectable option does not imply eligibility for it.

The age and interview-day separation among people living with HIV is the main threat to attributing the observed contrast to age itself. Recruitment-session arrangements and counselling exposure were not recorded sufficiently to resolve it. Participant-level site was also unavailable, preventing adjustment for differences between the two facilities. Clients attending these services are not representative of all potential users in Uganda. The cohort restriction from 251 to the parent sample of 232 preserves comparability between manuscripts but does not establish that excluded surplus records were ineligible. Recorded former and never PrEP users, and 32 prevention clients denying care at either named facility, differ from the protocol’s current-user eligibility wording. The care-restricted sensitivity analysis does not resolve the underlying recruitment discrepancy.

The benefit and concern summaries are questionnaire-derived scores rather than validated clinical measures. Their interpretation assumes that averaging the items is meaningful, and concurrent measurement with readiness leaves substantial scope for common-method effects and reverse direction of association. The exploratory factor models did not mutually adjust all factors and omitted the age-by-service interaction, so their estimates should not be described as independent predictors established across service groups. Some Poisson fitted means exceeded one, and expanded age comparisons are consequently reported using logistic standardisation. The logistic output does not provide standardised contrasts for each exploratory factor. Sensitivity models for immediate initiation, not-applicable responses and the care restriction used pooled age coefficients, not service-specific interaction contrasts.

The sample-size calculation addressed the parent willingness comparison rather than the present interaction or exploratory analyses. Small reluctance subgroups, multiple comparisons without multiplicity adjustment and 500-resample bootstrap intervals limit precision and increase the possibility of chance findings. Absence of a clear association is not evidence that a factor is irrelevant. Recall and social-desirability bias could affect self-reported information, finances and perceptions. Finally, the initiation item combines a fixed near-term preference with an information-dependent response; this useful distinction does not constitute a validated readiness scale or a commitment to initiate when a product becomes available.

## Conclusion

Nearly half of clients with applicable timing responses were ready to start injectable HIV treatment or prevention, while a similar proportion wanted further information about injectable options. Safety, appointment flexibility and privacy remained relevant even among ready respondents. Information sources, perceptions and financial circumstances were associated with readiness in exploratory analyses. HIV programmes should pair expanded product choice with clear counselling and accessible delivery arrangements, so that clients can make informed decisions and act on them when clinically appropriate.

## Data Availability

The participant-level data are subject to restricted access. Researchers should submit requests to Mildmay Research Centre Uganda at. Access requires approval from Mildmay Research Centre Uganda and a signed data-use agreement to protect participant confidentiality.

## Author Contributions

Conceptualization: Wandera Uthmaan Muluga, Drake Musoki, Chad D. Stecher.

Methodology: Stephen Charles Olet, Wandera Uthmaan Muluga, Drake Musoki, Chad D. Stecher, David Musoke.

Investigation: Wandera Uthmaan Muluga, Drake Musoki, Bernard Raymond Kihumuro, Farid Muluga, Ivan Ssebandeke, Fiona Among.

Data curation: Stephen Charles Olet, Wandera Uthmaan Muluga, Farid Muluga, Ivan Ssebandeke.

Formal analysis: Stephen Charles Olet.

Writing, original draft: Stephen Charles Olet.

Writing, review and editing: Stephen Charles Olet, Wandera Uthmaan Muluga, Drake Musoki, Bernard Raymond Kihumuro, Farid Muluga, Ivan Ssebandeke, Fiona Among, Chad D. Stecher, David Musoke.

Supervision: Wandera Uthmaan Muluga, Chad D. Stecher, David Musoke.

Funding acquisition: Wandera Uthmaan Muluga.

## Data Availability

Participant-level data are not publicly available because [confirmed ethical or legal restriction]. Researchers should direct data-access requests to Mildmay Research Centre Uganda at. Access requires approval from Mildmay Research Centre Uganda and a signed data-use agreement to protect participant confidentiality.

## Acknowledgments

We thank the survey participants and the clinical and research teams at Mildmay Hospital and Kajjansi Health Centre IV for their contribution to the parent study.

## Supporting information captions

S1 Appendix. Detailed analytical results and recruitment checks. Cohort flow, complete regression estimates, demographic and expanded logistic standardization, sensitivity analyses, multiple-response conditions, reasons for reluctance and aggregated recruitment metadata. Participant identifiers and private free text are excluded.

